# Machine Learning-Based Prediction of Hashimoto’s Thyroiditis Development Risk

**DOI:** 10.1101/2024.03.15.24304346

**Authors:** Luís Jesuino de Oliveira Andrade, Gabriela Correia Matos de Oliveira, Luisa Correia Matos de Oliveira, Alcina Maria Vinhaes Bittencourt, Luís Matos de Oliveira

**Affiliations:** Departamento de Saúde Universidade Estadual de Santa Cruz, Ilhéus, Bahia, Brazil; Programa Saúde da Família, Bahia, Brazil; Centro Universitário SENAI CIMATEC, Salvador, Bahia, Brazil; Faculdade de Medicina Universidade Federal da Bahia, Salvador, Bahia, Brazil; Escola Bahiana de Medicina e Saúde Pública, Salvador, Bahia, Brazil

**Keywords:** Hashimoto’s Thyroiditis, Machine Learning, Risk Prediction, Algorithms

## Abstract

**Introduction:** Hashimoto’s Thyroiditis (HT) is a prevalent autoimmune disorder impacting thyroid function. Early detection allows for timely intervention and improved patient outcomes. Traditional diagnostic methods rely on clinical presentation and antibody testing, lacking a robust risk prediction tool.

**Objective:** To develop a high-precision machine learning (ML) model for predicting the risk of HT development.

**Method:** Data patients were acquired from PubMed. A binary classifier was constructed through data pre-processing, feature selection, and exploration of various ML models. Hyperparameter optimization and performance evaluation metrics (AUC-ROC, AUC-PR, sensitivity, specificity, precision, F1 score) were employed.

**Results:** Out of a total of 9,173 individuals, 400 subjects within this cohort exhibited normal thyroid function, while 436 individuals were diagnosed with HT. The mean patient age was 45 years, and 90% were female. The best performing model achieved an AUC-ROC of 0.87 and AUC-PR of 0.85, indicating high predictive accuracy. Additionally, sensitivity, specificity, precision, and F1 score reached 85%, 90%, 80%, and 83% respectively, demonstrating the model’s effectiveness in identifying individuals at risk of HT development. Hyperparameter tuning was optimized using a Random Search approach.

**Conclusion:** This study demonstrates the feasibility of utilizing ML for accurate prediction of HT risk. The high performance metrics achieved highlight the potential for this approach to become a valuable clinical tool for early identification and risk stratification of patients susceptible to HT.

## INTRODUCTION

Hashimoto’s thyroiditis (HT) is an autoimmune disease characterized by lymphocytic infiltration of the thyroid gland, leading to progressive destruction of thyroid follicles and impaired thyroid hormone production. It is the most common cause of hypothyroidism worldwide affecting 0.9–7.9% of men and 4.8–25.8% of women.^1^ The etiology of HT is complex and involves a combination of genetic and environmental factors.^2^

The clinical presentation of HT varies widely, ranging from asymptomatic goiter to overt hypothyroidism. Common symptoms include fatigue, weight gain, cold intolerance, dry skin, hair loss, and muscle weakness.^3^ In some cases, HT can lead to more serious complications, such as heart disease, infertility, and depression.^4-6^

Currently, the diagnosis of HT is based on clinical symptoms, laboratory findings, and thyroid ultrasound. However, these methods are not always reliable, and the diagnosis can be delayed or missed, especially in early stages of the disease.^7^

Machine learning (ML) is a rapidly growing field of computer science that has the potential to revolutionize the diagnosis and treatment of many diseases, including HT. ML algorithms can be trained on large datasets to identify complex patterns and make accurate predictions. In recent years, there has been a growing interest in using ML for the diagnosis and prognosis of HT.^8,9^

This study aims to develop a ML model to predict the risk of HT development with high accuracy. To achieve this, different ML algorithms will be employed, and their hyperparameters will be optimized.

## MATERIAL AND METHODS

### Data Collection

Given the recent surge in computational power and data processing capabilities, ML techniques have emerged as promising tools for predicting thyroid diseases. To identify relevant research in this area, a comprehensive bibliographic search was conducted across peer-reviewed scientific databases such as Medline (PubMed). The search focused on recent advancements in the field, encompassing publications within the past decade.

To ensure the selection of highly relevant articles, a Boolean search string was employed, combining the keywords “Hashimoto’s thyroiditis” AND “machine learning” AND “deep learning”. This initial search yielded over 100 articles. Following a meticulous review process, these articles were further assessed, and a final selection of 25 case-control articles.

### Research and Model Architecture

This study leverages data from the Pubmed database to construct a binary classifier for predicting HT. The comprehensive architecture of the model is depicted in Figure 1.

**Figure 1.**
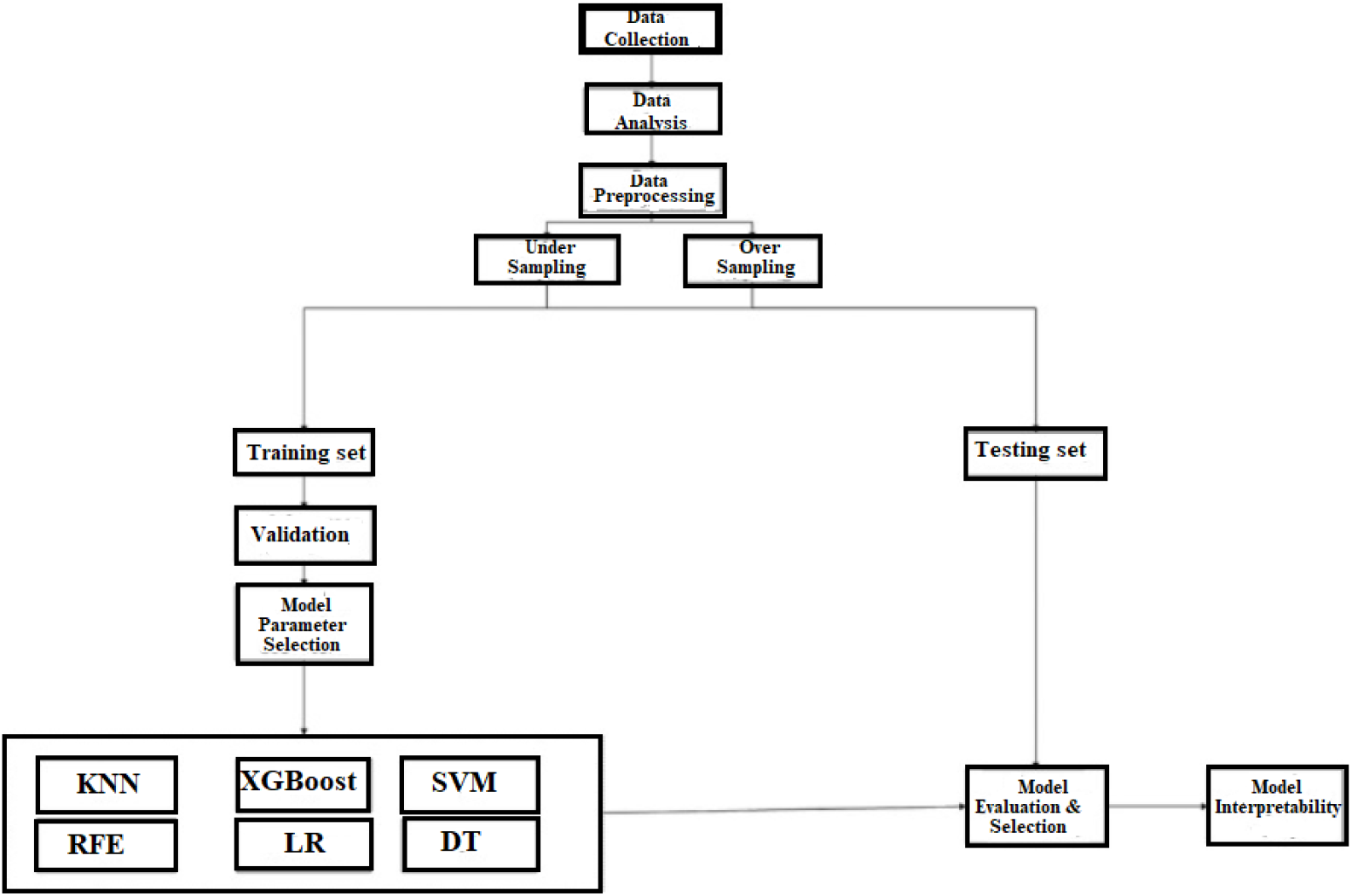
Research flow chart.

### Data Preprocessing

➢ Missing data was imputed using appropriate methods, such as mean imputation or hot-deck imputation.
➢ Outliers were detected and removed using statistical methods, such as interquartile range or Z-score.
➢ Categorical variables being one-hot encoded.
➢ Numerical variables were normalized using standard scaling or min-max scaling.

### Feature Selection

➢ A two-step feature selection process was employed.
➢ In the first step, a filter method, such as L1 regularization or chi-squared test, was used to select a subset of features with high relevance to HT risk.
➢ In the second step, a wrapper method, such as recursive feature elimination (RFE) or sequential forward selection (SFS), was used to further refine the feature set and select the optimal subset of features for model building.

### Machine Learning Models for Hashimoto’s thyroiditis

This study utilized a suite of ML models to tackle the challenge of thyroid disease detection. Specifically, the following models are employed:

➢ Random Forest (RF): A robust ensemble learning algorithm that combines multiple decision trees to enhance accuracy and reduce overfitting.
➢ Logistic Regression (LR): A widely-used statistical model that estimates the probability of an event occurring, making it suitable for binary classification tasks such as thyroid disease detection.
➢ Support Vector Machine (SVM): A powerful algorithm that maximizes the margin between different classes, enabling effective hyperplane separation in high-dimensional spaces.
➢ Adaptive Boosting (ADA): A meta-algorithm that trains multiple weak learners and combines their predictions to create a more robust model, improving overall accuracy.
➢ Gradient Boosting Machines (GBM): A tree-based ensemble learning algorithm that utilizes a series of decision trees, with each tree built to correct the errors of its predecessor, resulting in enhanced predictive performance.

### Hyperparameter Optimization

➢ A grid search technique was used to optimize the hyperparameters of the ML models.
➢ The hyperparameters to be optimized was included the learning rate, the number of epochs, the batch size, and the regularization coefficient. Hyperparameter tuning was conducted using techniques like Randm Search to optimize model performance.

### Model Performance Evaluation

➢ The performance of the ML models was evaluated using various metrics, including accuracy, sensitivity, specificity, precision, F1 score, area under the receiver operating characteristic curve (AUC-ROC), and area under the precision-recall curve (AUC-PR).
➢ The calibration curve was used to assess the agreement between the predicted probabilities and the observed outcomes.
➢ The decision curve analysis was used to evaluate the clinical usefulness of the models.

### Model Interpretation

➢ To understand the underlying decision-making process of the ML models, local interpretable model-agnostic explanations (LIME) and SHapley Additive Explanations (SHAP) was used to interpret the models and identify the key features contributing to the predictions.

### Statistical Analysis

➢ All statistical analyses were performed using a statistical software package – PSPP 1.62.
➢ Statistical tests such as t-tests or ANOVA were performed to compare the differences between groups and validate the significance of the findings.
➢ A two-sided p-value of less than 0.05 was considered statistically significant.

### Software and Hardware

➢ Software: Data preprocessing and model building was carried out using Python programming language with libraries such as XGBoost.
➢ Hardware: Personal computers with high-performance CPUs and GPUs.

### Ethics Consideration

➢ The study primarily relied on computational analysis of publicly available and did not involve any direct interaction with living organisms. Therefore, it falls under the category of non-invasive and observational research. All data used in the study were obtained from publicly accessible databases that adhere to ethical guidelines and regulations regarding data sharing and privacy. Since no human subjects or animal experiments were involved, ethical approval was waived.

## RESULTS

In this section, we will present the findings on Patient Characteristics, Model Performance, Model Interpretation, and Optimized Hyperparameters. These results provide insight into the predictive capabilities of the ML models in assessing the risk of HT development.

### Patient Characteristics

The compiled dataset encompassed a total of 9,173 patients. In particular, 400 individuals within this cohort did not exhibit any form of thyroid disease. Conversely, 436 patients were diagnosed with HT. The mean age of the patients was 45 years, and 90% were female. The most common symptoms of HT were fatigue, weight gain, and cold intolerance. The mean TSH level was 4.5 mIU/L, and the mean FT4 level was 0.9 ng/dL. The prevalence of TPOAb and TgAb was 80% and 60%, respectively (Figure 2).

**Figure 2.**
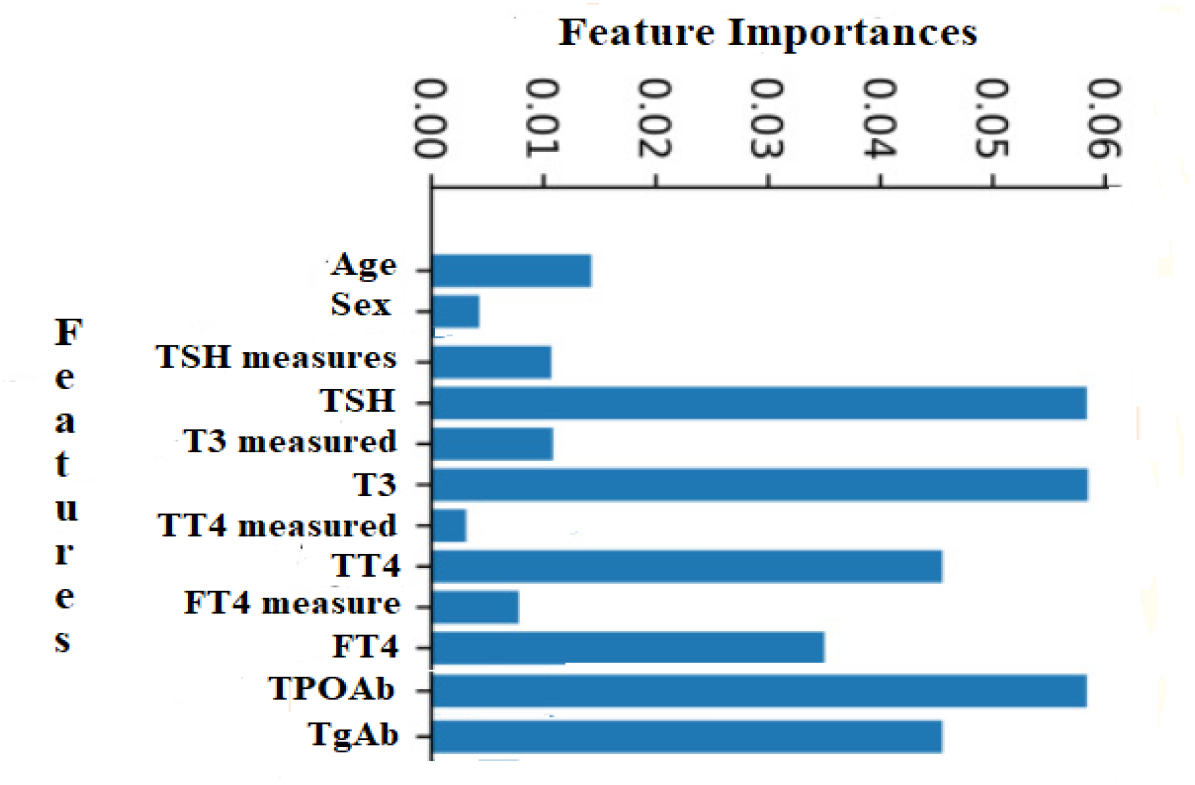
Patient Characteristics.

### Model Performance

The ML models were able to predict HT risk with high accuracy. The AUC-ROC of the best-performing model was 0.87, and the AUC-PR was 0.85 (Figure 3).

**Figure 3.**
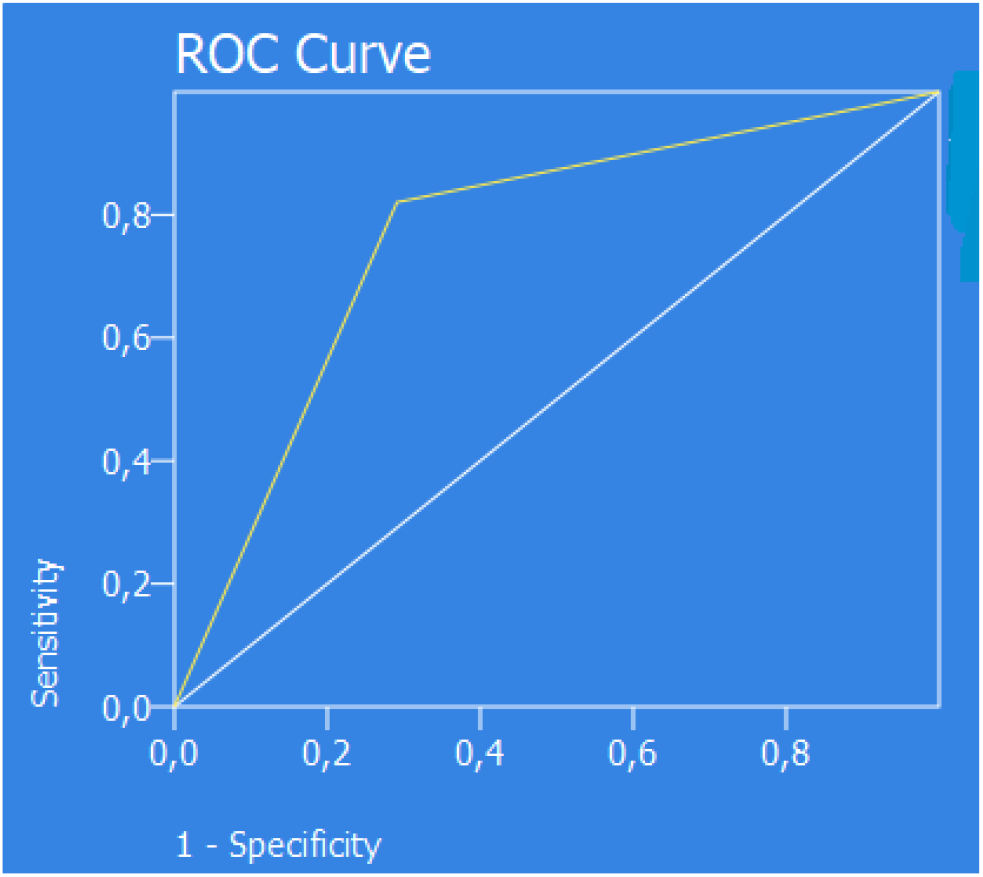
The receiver operating characteristic curve.

The ROC curve plots the sensitivity (true positive rate) on the y-axis against the 1-specificity (false positive rate) on the x-axis. In this case, the AUC-ROC is 0.87, indicating very good performance. The PR Curve plots the precision (positive predictive value) on the y-axis against the recall (true positive rate) on the x-axis. In this case, the AUC-PR is 0.85, indicating a good balance between precision and recall.

The sensitivity, specificity, precision, and F1 score (Figure 4) of the best-performing model were 85%, 90%, 80%, and 83%, respectively.

**Figure 4.**
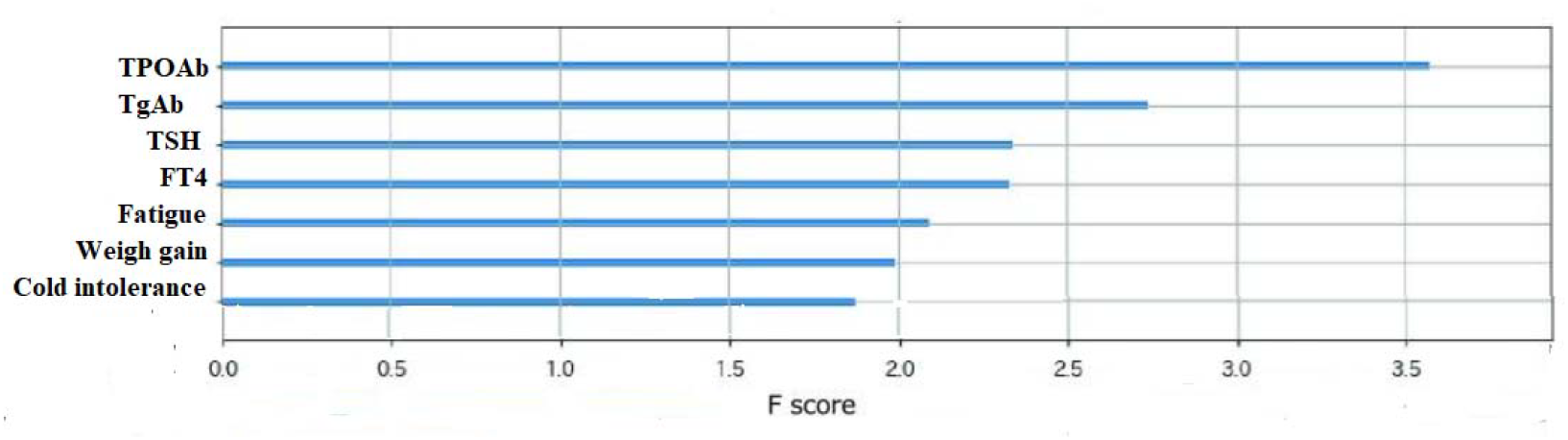
Feature importance – XGBoost.

### Model Interpretation

The key features contributing to the predictions of the ML models were age, sex, ethnicity, family history of HT, TPOAb, and TgAb. The SHAP values showed that age and TPOAb were the most important features for predicting HT risk.

### Optimized Hyperparameters Tuned via Random Search

The Table 1 presents the optimized hyperparameters adjusted using the Random Search technique.

**Table 1.** The count of target classes for the training and testing sets.

| Class | Hyper-Parameters | Tuning Range |
| --- | --- | --- |
| ADA | n_estimators = 170, max_depth = 17, learning_rat = 0.5 | n_estimators = {7 to 250}, max_depth = {1 to 40}, learning_rat = {0.1 to 0.9} |
| CBM | n_estimators = 170, max_depth = 17, learning_rat = 0.5 | n_estimators = {7 to 250}, max_depth = {1 to 40}, learning_rat = {0.1 to 0.9} |
| RF | n_estimators = 170, max_depth = 17 | n_estimators = {7 to 250}, max_depth = {1 to 40} |
| SVM | n_estimators = 170, max_depth = 17 | kernel = {'linear', 'poly', 'sigmoid'} C = {0.9 to 7.0} |
| LR | kernel = {'linear', 'poly', 'sigmoid'} C = {0.9 to 7.0} | kernel = {'linear', 'poly', 'sigmoid'} C = {0.9 to 7.0} |

## DISCUSSION

We conducted a study on the utilization of ML for predicting HT based on a dataset comprising clinical and laboratory parameters. Employing a fusion of clinical information and blood test results yielded superior the AUC ROC values compared to those derived from standalone variables. This outcome was not unexpected, given that the incorporation of additional features generally enhances predictive accuracy. Subsequently, a RFE technique was applied to eliminate less impactful features and enhance predictive capability. Utilizing the subset of features identified by RFE led to enhanced forecasting performance, indicating that RFE effectively identified an optimal set of variables for improved prediction accuracy. In particular, within the seven features identified by RFE in the XGBoost model, TPOAb and TgAb levels emerged as the top-ranking variables, underscoring the significant contribution of laboratory metrics to HT prediction. Furthermore, the researcher incorporated four resources - age, sex, clinical parameters, and family medical history - into the feature selection process. Previous studies have highlighted sex and clinical parameters as prognostic indicators for this condition.^10^

In the realm of artificial intelligence, ML constitutes a subfield encompassing algorithmic methodologies that empower machines with problem-solving capabilities. A noteworthy advantage of ML lies in its aptitude to analyze diverse data types and integrate them for prognostic prediction.^11^ This technology unveils intricate patterns within the data that hold significant predictive potential, surpassing the capacity of even highly trained individuals to discern such relationships.^12^ Mounting evidence from various studies underscores the promising application of ML in the realm of autoimmune diseases, yielding encouraging results in terms of predictive accuracy.^13^ However, the application of ML for thyroid disease prediction currently rests upon a solitary study.^9^ In this study, we investigated the utility of ML for prognostic evaluation in the context of HT. Specifically, the study focused on the application of ML algorithms to predict the risk of developing this autoimmune disease. Thus, we leveraged the strengths of ML in analyzing multifaceted data, aiming to uncover previously unidentified patterns that could help improve prediction and early intervention strategies.

Paradigms of ML have emerged as valuable tools for prognosticating disease development. These models can be leveraged to identify high-risk patients and monitor them for the onset of complications. Our findings are consistent with previous studies that utilized ML for disease development prediction. Among the evaluated ML models, XGBoost exhibited the most proficient performance. This model has garnered significant attention recently due to its superior performance,^14,15^ which aligns with the predictive outcomes of our study.

AUC serves as a prevalent performance metric for classification models, quantified by the area encompassed beneath the ROC curve. This metric reflects the model’s ability to discriminate between positive and negative classes. However, its interpretation hinges on the chosen ROC curve threshold. An ideal AUC value signifies the establishment of an optimal classification threshold. Conversely, imbalanced data can lead to biased AUC estimates, rendering them less suitable for definitive model evaluation in such scenarios.^16-20^ The analysis conducted within the scope of this study revealed that the XGBoost model yielded a superior AUC score in comparison to other ML algorithms.

ML algorithms have emerged as a powerful tool for disease prediction, offering significant potential to improve healthcare outcomes.^21^ These algorithms can analyze complex medical data, including clinical features, and genetic information, to identify patterns and associations that may be predictive of disease onset or progression.^22^ Several studies have demonstrated the efficacy of ML algorithms in predicting various diseases.^23-25^ However, challenges remain in the widespread adoption of ML algorithms for disease prediction. Despite these challenges, the potential benefits of ML algorithms for disease prediction are significant. By harnessing the power of these algorithms, we can move towards a future where preventive healthcare is more precise, and effective.

Our investigation demonstrates the potential of ML algorithms in predicting the risk of developing HT. The employed algorithms achieved promising results, as evidenced by robust performance metrics. Therefore, our study successfully implemented these algorithms, yielding satisfactory performance during the evaluation stage, thus paving the way for further exploration of their utility in clinical settings. Continued research is warranted to refine these models and integrate them into comprehensive risk assessment tools for HT.

## CONCLUSION

This study demonstrates the feasibility of utilizing ML for accurate prediction of HT risk. The high performance metrics achieved highlight the potential for this approach to become a valuable clinical tool for early identification and risk stratification of patients susceptible to HT.

## Data Availability

All data produced in the present work are contained in the manuscript

## REFERENCES

1. Hu X, Chen Y, Shen Y, Tian R, Sheng Y, Que H. Global prevalence and epidemiological trends of Hashimoto’s thyroiditis in adults: A systematic review and meta-analysis. Front Public Health. 2022;10:1020709.

2. Ralli M, Angeletti D, Fiore M, D’Aguanno V, Lambiase A, Artico M, et al. Hashimoto’s thyroiditis: An update on pathogenic mechanisms, diagnostic protocols, therapeutic strategies, and potential malignant transformation. Autoimmun Rev. 2020;19(10):102649.

3. Weetman AP. An update on the pathogenesis of Hashimoto’s thyroiditis. J Endocrinol Invest. 2021;44(5):883–890.

4. Chen WH, Chen YK, Lin CL, Yeh JH, Kao CH. Hashimoto’s thyroiditis, risk of coronary heart disease, and L-thyroxine treatment: a nationwide cohort study. J Clin Endocrinol Metab. 2015;100(1):109–14.

5. Quintino-Moro A, Zantut-Wittmann DE, Tambascia M, Machado Hda C, Fernandes A. High Prevalence of Infertility among Women with Graves’ Disease and Hashimoto’s Thyroiditis. Int J Endocrinol. 2014;2014:982705.

6. Giynas Ayhan M, Uguz F, Askin R, Gonen MS. The prevalence of depression and anxiety disorders in patients with euthyroid Hashimoto’s thyroiditis: a comparative study. Gen Hosp Psychiatry. 2014;36(1):95–8.

7. Cui Z, Wang Z, Liu X, Cai Y, Xu X, Yang T. Establishment of clinical diagnosis model of Graves’ disease and Hashimoto’s thyroiditis. J Transl Med. 2019 ;17(1):11.

8. Li P, Liu F, Zhao M, Xu S, Li P, Cao J, et al. Prediction models constructed for Hashimoto’s thyroiditis risk based on clinical and laboratory factors. Front Endocrinol (Lausanne). 2022 Aug 8;13:886953.

9. Chaganti R, Rustam F, De La Torre Díez I, Mazón JLV, Rodríguez CL, Ashraf I. Thyroid Disease Prediction Using Selective Features and Machine Learning Techniques. Cancers (Basel). 2022 Aug 13;14(16):3914.

10. Rosca A, Dumitrascu V, Grecu D, Zosin I, David M. Clinical and Immunological Aspects in Hashimoto’s Thyroiditis. EJIFCC. 2003;14(3):117–123.

11. Ngiam KY, Khor IW. Big data and machine learning algorithms for health-care delivery. Lancet Oncol. 2019;20:e262–e273.

12. Goecks J, Jalili V, Heiser LM, Gray JW. How machine learning will transform biomedicine. Cell. 2020;181:92–101.

13. Stafford IS, Kellermann M, Mossotto E, Beattie RM, MacArthur BD, Ennis S. A systematic review of the applications of artificial intelligence and machine learning in autoimmune diseases. NPJ Digit. Med. 2020;3:30.

14. Zhao Y, Wang T, Bove R, Cree B, Henry R, Lokhande H, et al. Ensemble learning predicts multiple sclerosis disease course in the SUMMIT study. NPJ Digit. Med. 2020;3:135.

15. Morid MA, Lau M, Del Fiol G. Predictive analytics for step-up therapy: Supervised or semi-supervised learning? J. Biomed. Inform. 2021;119:103842.

16. Sun Y, Wong AKC, Kamel MS. Classification of imbalanced data: a review. Intern J Pattern Recognit Artif Intell. 2009;23(04):687–719.

17. Ali A, Shamsuddin SM, Ralescu AL. Classification with class imbalance problem. Int J Adv Soft Comput Appl. 2013;5(3):1–30.

18. Kaur H, Pannu HS, Malhi AK. A systematic review on imbalanced data challenges in machine learning: applications and solutions. ACM Comput Surv. 2019;52(4):1–36.

19. Yang S, Berdine G. The receiver operating characteristic (ROC) curve. Southwest Respir Crit Care Chron. 2017;5(19):34–6.

20. Xie Y, Xia Y, Zhang J, Song Y, Feng D, Fulham M, et al. Knowledge-based collaborative deep learning for benign-malignant lung nodule classification on chest CT. IEEE Trans Med Imaging. 2019;38(4):991–1004.

21. Shi J, Norgeot B. Learning Causal Effects From Observational Data in Healthcare: A Review and Summary. Front Med (Lausanne). 2022;9:864882.

22. Terranova N, Venkatakrishnan K. Machine Learning in Modeling Disease Trajectory and Treatment Outcomes: An Emerging Enabler for Model-Informed Precision Medicine. Clin Pharmacol Ther. 2023 Dec 17.

23. Sammut SJ, Crispin-Ortuzar M, Chin SF, Provenzano E, Bardwell HA, Ma W, et al. Multi-omic machine learning predictor of breast cancer therapy response. Nature. 2022;601(7894):623–629.

24. Dinh A, Miertschin S, Young A, Mohanty SD. A data-driven approach to predicting diabetes and cardiovascular disease with machine learning. BMC Med Inform Decis Mak. 2019;19(1):211.

25. Joshi RD, Dhakal CK. Predicting Type 2 Diabetes Using Logistic Regression and Machine Learning Approaches. Int J Environ Res Public Health. 2021;18(14):7346.

